# Natural History, Phenotype Spectrum and Clinical Outcomes of Desmin (*DES*)-Associated Cardiomyopathy

**DOI:** 10.1101/2024.08.24.24311904

**Authors:** Babken Asatryan, Marina Rieder, Brittney Murray, Steven A. Muller, Crystal Tichnell, Alessio Gasperetti, Richard T. Carrick, Emily Joseph, Doris G. Leung, Anneline S.J.M. te Riele, Stefan L. Zimmerman, Hugh Calkins, Cynthia A. James, Andreas S. Barth

## Abstract

**Background:** Pathogenic/likely pathogenic (P/LP) desmin (*DES*) variants cause heterogeneous cardiomyopathy and/or skeletal myopathy phenotypes. Limited data suggest a high incidence of major adverse cardiac events (MACE), including cardiac conduction disease (CCD), sustained ventricular arrhythmias (VA), and heart failure (HF) events (HF hospitalization, LVAD/cardiac transplant, HF-related death), in patients with P/LP *DES* variants. However, pleiotropic presentation and small cohort sizes have limited clinical phenotype and outcome characterization.

**Objectives:** We aimed to describe the natural history, phenotype spectrum, familial penetrance and outcomes in patients with P/LP *DES* variants through a systematic review and individual patient data meta-analysis using published reports.

**Methods:** We searched Medline (PubMed) and Embase for studies that evaluated cardiac phenotypes in patients with P/LP *DES* variants. Cardiomyopathy diagnosis or occurrence of MACE were considered evidence of cardiac involvement/penetrance. Lifetime event-free survival from CCD, sustained VA, HF events, and composite MACE was assessed.

**Results:** Out of 4,212 screened publications, 71 met the inclusion criteria. A total of 230 patients were included (52.6% male, 52.2% probands, median age: 31 years [22.0; 42.8] at first evaluation, median follow-up: 3 years [0; 11.0]). Overall, 124 (53.9%) patients were diagnosed with cardiomyopathy, predominantly dilated cardiomyopathy (14.8%), followed by restrictive cardiomyopathy (13.5%), whereas other forms were less common: arrhythmogenic cardiomyopathy (7.0%), hypertrophic cardiomyopathy (6.1%), arrhythmogenic right ventricular cardiomyopathy (5.2%), and other forms (7.4%). Overall, 132 (57.4%) patients developed MACE, with 96 [41.7%] having CCD, 36 [15.7%] sustained VA, and 43 [18.7%] HF events. Familial penetrance of cardiac disease was 63.6% among relatives with P/LP *DES* variants. Male sex was associated with increased risk of sustained VA (HR 2.28, p=0.02) and HF events (HR 2.45, p=0.008).

**Conclusions:** *DES* cardiomyopathy exhibits heterogeneous phenotypes and distinct natural history, characterized by high familial penetrance and substantial MACE burden. Male patients face higher risk of sustained VA events.

## INTRODUCTION

Desmin, encoded by the *DES* gene, is a muscle-specific type III intermediate filament highly expressed in cardiac tissue, particularly in the Purkinje fibers,(1,2), as well as in skeletal and smooth muscle cells. Desmin interacts with various cellular structures, such as the intercalated disk through desmoplakin, sarcomeric Z-discs, and the nuclear envelope protein lamin A/C, forming a complex three-dimensional intracellular network essential for the transmission of cellular mechanical force and mechanochemical sensing.(2,3) Pathogenic (P) or likely pathogenic (LP) *DES* variants can lead to desminopathies, a spectrum of cardiomyopathy or skeletal myopathy phenotypes, either presenting in isolation or in combination.(4) Small cohort studies suggest marked inter- and intrafamilial heterogeneity in phenotype in patients with P/LP *DES* variants spanning across the morpho-functional spectrum of cardiomyopathies. Sustained ventricular arrhythmias (VA), advanced or complete atrioventricular conduction disease, and heart failure (HF) events, have been reported in patients with *DES-*associated cardiomyopathy,(5) but the burden of such events in this population remains unknown. While these features suggest that *DES*-associated cardiomyopathy may require a tailored approach as a distinct form of cardiomyopathy, the genetic landscape, natural history, and prognosis of *DES* cardiomyopathy are largely unexplored, and consequently, the management of affected families remains poorly defined.

Given the distinct disease mechanisms, phenotypic expressions and outcomes observed in different genetic forms of cardiomyopathies,(6–12) a better understanding of *DES*-associated cardiomyopathy is crucial for developing precision management strategies.(13,14) However, assembly of large cohorts of such patients is challenged by the rare nature of *DES*-associated disease.(15,16) Moreover, *DES* disease presents in both adult-onset and childhood-onset forms, with clinical manifestations ranging from cardiomyopathy and/or skeletal myopathy to multisystem disease with involvement of cardiac, respiratory, bulbar, facial, and smooth muscles.(17) This pleiotropy results in patients presenting across a wide array of clinical contexts and disciplines with a plethora of symptoms,(4,18) complicating the assessment of the full spectrum of cardiac disease expressions associated with *DES* disease. Therefore, an integrated approach is necessary to provide a holistic understanding of *DES-*associated cardiomyopathy.

As a first step to overcome these challenges, we conducted a systematic review and individual patient data meta-analysis of patients harboring P/LP *DES* variants to thoroughly characterize the genetic landscape, natural history, phenotype spectrum, familial penetrance and clinical outcomes in patients with P/LP *DES* variants identified via clinical genetic testing.

## METHODS

### Compliance with Ethical Standards

The data used in the systematic review and individual patient data meta-analysis are publicly available and ethics approval was not required. The dataset generated through the extraction of data from published studies will be made available upon reasonable request.

### Data Source and Search Strategy

The systematic review and individual patient data meta-analysis were conducted following the recommendations of the Preferred Reporting Items for Systematic Reviews and Meta-analysis (PRISMA) statement.(19) With the assistance of an experienced medical research librarian (EJ), we established a systematic search strategy. All searches were run in the following databases: Medline (PubMed) and Embase to identify citations published from 1998 (when the first P/LP *DES* variants were reported in association with disease)(20) until May 31, 2024. In Embase, conference abstracts were excluded. For the search strategies designed for Medline (PubMed) and Embase, controlled vocabulary terms for each concept were identified and combined with keyword synonyms (see **Table S1** for complete search strings).

An initial screening of the title and abstract of each study were conducted by two independent investigators (BA and MR) using Covidence to identify studies potentially including human data on patients harboring *DES* variants. Subsequently, eligible papers underwent full-text assessment to confirm that they reported cases meeting the patient inclusion criteria. Disagreements on inclusion were discussed until consensus was reached between both investigators. References of included articles and published review articles were also manually screened for additional pertinent articles. If the same patient was presumed to be reported by more than one studies (based on variant specifics: same base change and amino acid change; sex, age at presentation/evaluation, also with respect to date of publication, center of origin [for single center studies], and corresponding authors), only the one containing more clinical phenotype and outcome description was included. In case pertinent data related to a case was reported in different publications, data was merged using unique case identifiers.

### Patient population—inclusion and exclusion criteria

Patients were included in the study if they met the following criteria:

i. harbored a P/LP variant in *DES*, as determined at core lab adjudication;(21)
ii. were alive at the time of the first evaluation; and
iii. had at least one clinical encounter with cardiac evaluation (at least ECG and either echocardiogram or cardiac magnetic resonance imaging [CMR]).

Patients were excluded from the study if they met any of the following criteria:

i. had a confirmed or probable digenic inheritance (i.e. carried additional variant(s) of uncertain significance [VUS], LP or P variant in another gene implicated in inherited arrhythmias or cardiomyopathies); or
ii. experienced a major adverse cardiac event (MACE) due to a presumed alternative cause (e.g. ventricular fibrillation in the context of acute myocardial infarction).

### Data Extraction

Study details, detailed individual-level demographics, clinical presentation, symptoms, clinical evaluation findings, genetic information, family history, as well as specific outcomes, were extracted through the review of published manuscripts by one investigator (BA). NM_001927.4 was accepted as the reference transcript. All *DES* variants were adjudicated in accordance with the standards and guidelines set forth by the American College of Medical Genetics and Genomics (ACMG) and the Association for Molecular Pathology (AMP).(21) Variants fulfilling criteria for pathogenicity only at compound heterozygous or homozygous state were reported as such. Variants were considered as inherited if the biological mother and/or the father was heterozygous for the P/LP *DES* variant; *de novo* origin was determined as confirmed when both parents were non-carriers of the variant; and the variant was classified as of unknown origin if both parental samples were unavailable, or when the only available parental sample did not carry it.

### Data Synthesis and Outcomes

The primary outcomes were the development of 1) cardiac conduction disease (CCD) requiring cardiac implantable electronic device (CIED) implantation, 2) sustained VA, 3) HF events, and 4) MACE (defined as a composite of CCD, sustained VA and HF events). Secondary outcomes included the development of a) cardiovascular death, and b) all-cause death. Sustained VA was defined as a composite of the occurrence of sustained ventricular tachycardia (VT), appropriate ICD interventions, ventricular fibrillation/flutter (VF), and sudden cardiac arrest (SCA) / sudden cardiac death (SCD) episodes, as reported. CCD events were defined as II-degree Mobitz type II, advanced degree or complete AV block, any symptomatic AV block or other CCD that led to CIED implantation. HF events were defined as a composite of hospitalization for HF, left ventricular assist device (LVAD) implantation/cardiac transplant, and HF-related death. As prognosis assessment was exclusively based on hard endpoints, both cross-sectional and longitudinal data were used for outcome analysis. Time to event was defined as the time from birth to first incident outcome of interest or censoring by last clinical encounter.

For patients with multiple clinical encounters, phenotypes at both presentation and last follow-up were recorded, while those with a single encounter had the ascertained phenotype considered as both initial and final observed phenotype. Penetrance was defined as the prevalence of disease in patients with a P/LP *DES* variant, i.e. the percentage of relatives with a P/LP *DES* variant with a cardiomyopathy phenotype and/or MACE (combined cardiomyopathy/MACE penetrance). The complete list of all included studies is provided in **Table S2**.

### Statistical Analysis

Continuous, normally distributed variables were summarized using mean (SD) and compared using 2-sample Student’s t-tests. Non-normally distributed continuous data were summarized using median and interquartile range and compared using Wilcoxon-rank sum tests. Categorical data were summarized using percentages and compared using χ2 or Fisher exact tests, as appropriate. Time-to-event analysis was performed using Cox proportional hazard regression using age as the timescale and birth as time zero. Survival duration was calculated from birth to date of censoring. Hazard ratios (HRs) are reported with 95% confidence intervals (CIs). α was set at 0.05 for statistical significance. Sex was a prespecified variable for analysis. Data were analyzed using R statistical computing, using the *survival, survminer* and *ggplot2* packages.(22)

## RESULTS

### Demographic and Clinical Characteristics

Out of 4,212 screened records, 71 met inclusion criteria and were subsequently included in the study. The complete study flowchart for identification, screening, and eligibility assessment of studies is displayed in **Figure S1**. Overall, 230 patients with 74 unique P/LP *DES* variants were included and followed for a median of 3.0 years (0.0; 11.0). Of them, 121 (52.6%) were male, and 120 (52.2%) were probands (**Table 1**). In the overall population, median age was 31.0 [22.0; 42.8] years at first evaluation, and 27 (22.5%) probands had their first evaluation prior to age 18. Out of 120 probands, 24 (20.0%) had a family history of a cardiomyopathy, 30 (25.0%) had a first- or a second-degree relative with a CIED, 15 (12.5%) had a family member who died of SCD, and 6 (5.0%) had a family history of cardiac transplant. Skeletal muscle weakness was the most common reason for first clinical evaluation (n=72, 31.3%), followed by familial evaluation (n=43, 18.7%) and syncope (n=35, 15.2%).

**Table 1.**
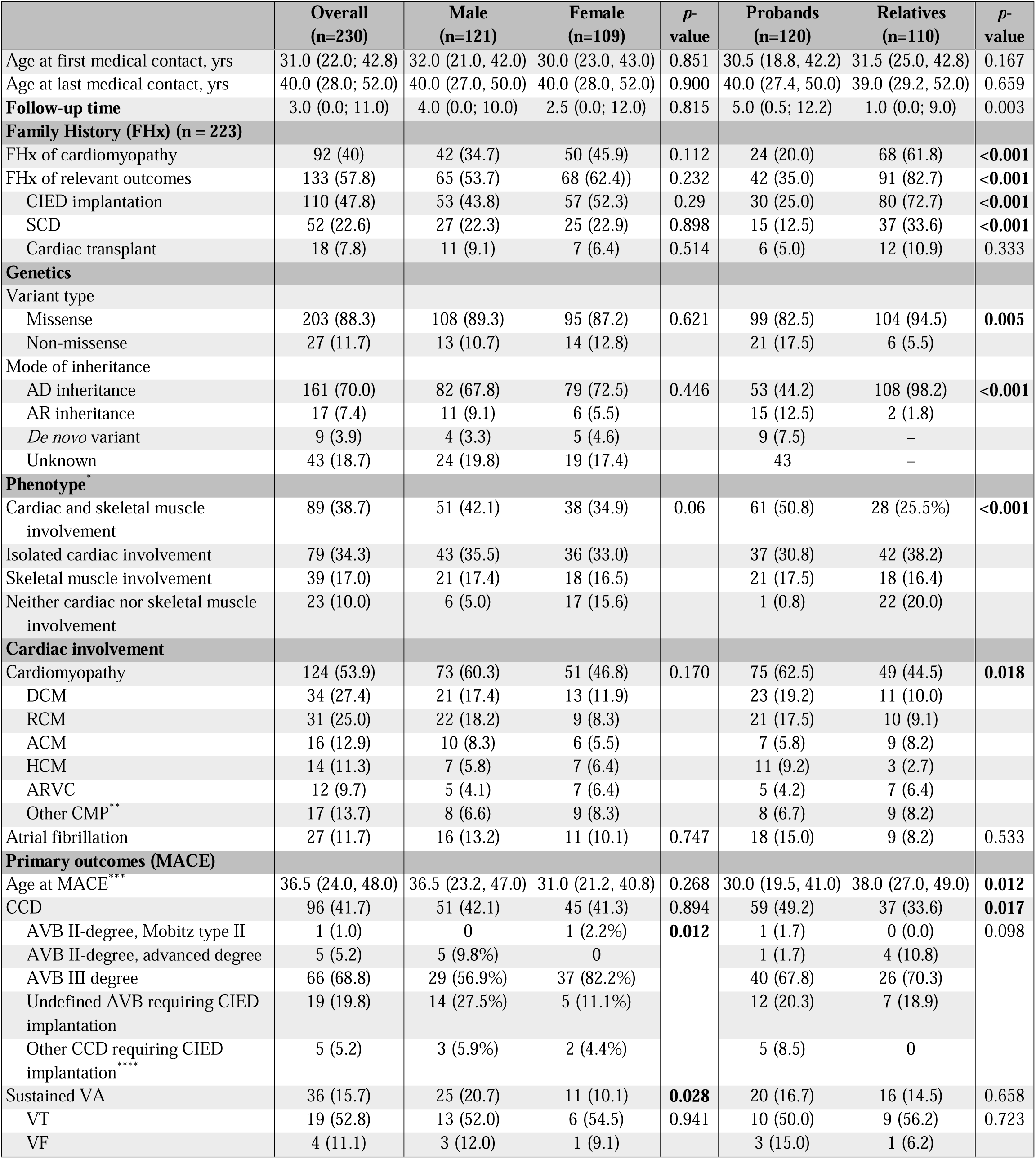
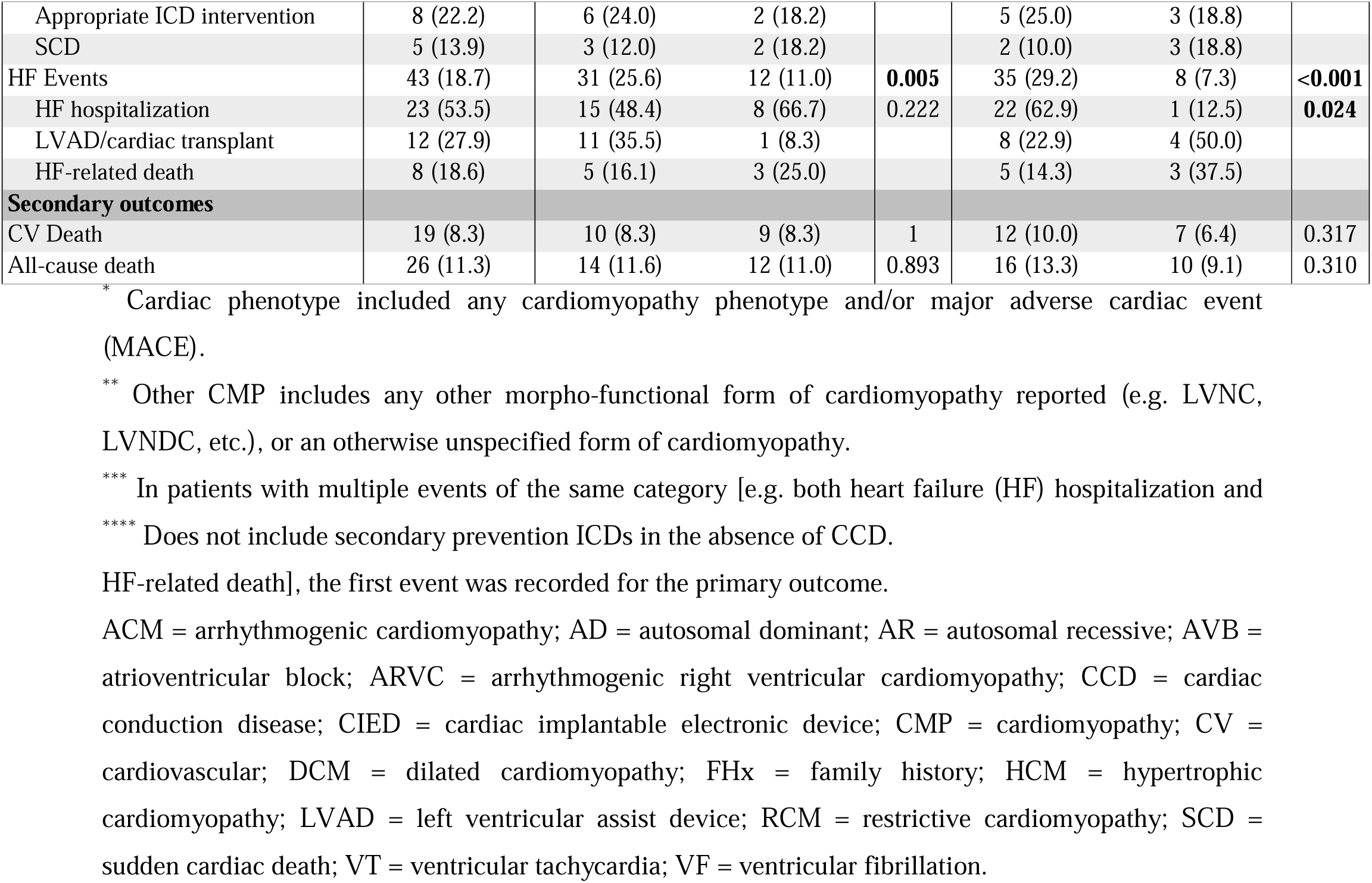
Demographic and clinical characteristics of the study population.

|  | Overall<br>(n=230) | Male<br>(n=121) | Female<br>(n=109) | p-<br>value | Probands<br>(n=120) | Relatives<br>(n=110) | p-<br>value |
| --- | --- | --- | --- | --- | --- | --- | --- |
| Age at first medical contact, yrs | 31.0 (22.0; 42.8) | 32.0 (21.0, 42.0) | 30.0 (23.0, 43.0) | 0.851 | 30.5 (18.8, 42.2) | 31.5 (25.0, 42.8) | 0.167 |
| Age at last medical contact, yrs | 40.0 (28.0; 52.0) | 40.0 (27.0, 50.0) | 40.0 (28.0, 52.0) | 0.900 | 40.0 (27.4, 50.0) | 39.0 (29.2, 52.0) | 0.659 |
| **Follow-up time** | 3.0 (0.0; 11.0) | 4.0 (0.0; 10.0) | 2.5 (0.0; 12.0) | 0.815 | 5.0 (0.5; 12.2) | 1.0 (0.0; 9.0) | 0.003 |
| **Family History (FHx) (n = 223)** |  |  |  |  |  |  |  |
| FHx of cardiomyopathy | 92 (40) | 42 (34.7) | 50 (45.9) | 0.112 | 24 (20.0) | 68 (61.8) | <0.001 |
| FHx of relevant outcomes | 133 (57.8) | 65 (53.7) | 68 (62.4) | 0.232 | 42 (35.0) | 91 (82.7) | <0.001 |
| CIED implantation | 110 (47.8) | 53 (43.8) | 57 (52.3) | 0.29 | 30 (25.0) | 80 (72.7) | <0.001 |
| SCD | 52 (22.6) | 27 (22.3) | 25 (22.9) | 0.898 | 15 (12.5) | 37 (33.6) | <0.001 |
| Cardiac transplant | 18 (7.8) | 11 (9.1) | 7 (6.4) | 0.514 | 6 (5.0) | 12 (10.9) | 0.333 |
| **Genetics** |  |  |  |  |  |  |  |
| Variant type |  |  |  |  |  |  |  |
| Missense | 203 (88.3) | 108 (89.3) | 95 (87.2) | 0.621 | 99 (82.5) | 104 (94.5) | 0.005 |
| Non-missense | 27 (11.7) | 13 (10.7) | 14 (12.8) |  | 21 (17.5) | 6 (5.5) |  |
| Mode of inheritance |  |  |  |  |  |  |  |
| AD inheritance | 161 (70.0) | 82 (67.8) | 79 (72.5) | 0.446 | 53 (44.2) | 108 (98.2) | <0.001 |
| AR inheritance | 17 (7.4) | 11 (9.1) | 6 (5.5) |  | 15 (12.5) | 2 (1.8) |  |
| De novo variant | 9 (3.9) | 4 (3.3) | 5 (4.6) |  | 9 (7.5) | – |  |
| Unknown | 43 (18.7) | 24 (19.8) | 19 (17.4) |  | 43 | – |  |
| **Phenotype\*** |  |  |  |  |  |  |  |
| Cardiac and skeletal muscle involvement | 89 (38.7) | 51 (42.1) | 38 (34.9) | 0.06 | 61 (50.8) | 28 (25.5%) | <0.001 |
| Isolated cardiac involvement | 79 (34.3) | 43 (35.5) | 36 (33.0) |  | 37 (30.8) | 42 (38.2) |  |
| Skeletal muscle involvement | 39 (17.0) | 21 (17.4) | 18 (16.5) |  | 21 (17.5) | 18 (16.4) |  |
| Neither cardiac nor skeletal muscle involvement | 23 (10.0) | 6 (5.0) | 17 (15.6) |  | 1 (0.8) | 22 (20.0) |  |
| **Cardiac involvement** |  |  |  |  |  |  |  |
| Cardiomyopathy | 124 (53.9) | 73 (60.3) | 51 (46.8) | 0.170 | 75 (62.5) | 49 (44.5) | 0.018 |
| DCM | 34 (27.4) | 21 (17.4) | 13 (11.9) |  | 23 (19.2) | 11 (10.0) |  |
| RCM | 31 (25.0) | 22 (18.2) | 9 (8.3) |  | 21 (17.5) | 10 (9.1) |  |
| ACM | 16 (12.9) | 10 (8.3) | 6 (5.5) |  | 7 (5.8) | 9 (8.2) |  |
| HCM | 14 (11.3) | 7 (5.8) | 7 (6.4) |  | 11 (9.2) | 3 (2.7) |  |
| ARVC | 12 (9.7) | 5 (4.1) | 7 (6.4) |  | 5 (4.2) | 7 (6.4) |  |
| Other CMP** | 17 (13.7) | 8 (6.6) | 9 (8.3) |  | 8 (6.7) | 9 (8.2) |  |
| Atrial fibrillation | 27 (11.7) | 16 (13.2) | 11 (10.1) | 0.747 | 18 (15.0) | 9 (8.2) | 0.533 |
| **Primary outcomes (MACE)** |  |  |  |  |  |  |  |
| Age at MACE*** | 36.5 (24.0, 48.0) | 36.5 (23.2, 47.0) | 31.0 (21.2, 40.8) | 0.268 | 30.0 (19.5, 41.0) | 38.0 (27.0, 49.0) | 0.012 |
| CCD | 96 (41.7) | 51 (42.1) | 45 (41.3) | 0.894 | 59 (49.2) | 37 (33.6) | 0.017 |
| AVB II-degree, Mobitz type II | 1 (1.0) | 0 | 1 (2.2%) | 0.012 | 1 (1.7) | 0 (0.0) | 0.098 |
| AVB II-degree, advanced degree | 5 (5.2) | 5 (9.8%) | 0 |  | 1 (1.7) | 4 (10.8) |  |
| AVB III degree | 66 (68.8) | 29 (56.9%) | 37 (82.2%) |  | 40 (67.8) | 26 (70.3) |  |
| Undefined AVB requiring CIED implantation | 19 (19.8) | 14 (27.5%) | 5 (11.1%) |  | 12 (20.3) | 7 (18.9) |  |
| Other CCD requiring CIED implantation**** | 5 (5.2) | 3 (5.9%) | 2 (4.4%) |  | 5 (8.5) | 0 |  |
| Sustained VA | 36 (15.7) | 25 (20.7) | 11 (10.1) | 0.028 | 20 (16.7) | 16 (14.5) | 0.658 |
| VT | 19 (52.8) | 13 (52.0) | 6 (54.5) | 0.941 | 10 (50.0) | 9 (56.2) | 0.723 |
| VF | 4 (11.1) | 3 (12.0) | 1 (9.1) |  | 3 (15.0) | 1 (6.2) |  |
| Appropriate ICD intervention | 8 (22.2) | 6 (24.0) | 2 (18.2) |  | 5 (25.0) | 3 (18.8) |  |
| SCD | 5 (13.9) | 3 (12.0) | 2 (18.2) |  | 2 (10.0) | 3 (18.8) |  |
| HF Events | 43 (18.7) | 31 (25.6) | 12 (11.0) | **0.005** | 35 (29.2) | 8 (7.3) | **<0.001** |
| HF hospitalization | 23 (53.5) | 15 (48.4) | 8 (66.7) | 0.222 | 22 (62.9) | 1 (12.5) | **0.024** |
| LVAD/cardiac transplant | 12 (27.9) | 11 (35.5) | 1 (8.3) |  | 8 (22.9) | 4 (50.0) |  |
| HF-related death | 8 (18.6) | 5 (16.1) | 3 (25.0) |  | 5 (14.3) | 3 (37.5) |  |
| **Secondary outcomes** |  |  |  |  |  |  |  |
| CV Death | 19 (8.3) | 10 (8.3) | 9 (8.3) | 1 | 12 (10.0) | 7 (6.4) | 0.317 |
| All-cause death | 26 (11.3) | 14 (11.6) | 12 (11.0) | 0.893 | 16 (13.3) | 10 (9.1) | 0.310 |

\* Cardiac phenotype included any cardiomyopathy phenotype and/or major adverse cardiac event (MACE).
\*\* Other CMP includes any other morpho-functional form of cardiomyopathy reported (e.g. LVNC, LVNDC, etc.), or an otherwise unspecified form of cardiomyopathy.
\*\*\* In patients with multiple events of the same category [e.g. both heart failure (HF) hospitalization and
\*\*\*\* Does not include secondary prevention ICDs in the absence of CCD.
HF-related death], the first event was recorded for the primary outcome.
ACM = arrhythmogenic cardiomyopathy; AD = autosomal dominant; AR = autosomal recessive; AVB = atrioventricular block; ARVC = arrhythmogenic right ventricular cardiomyopathy; CCD = cardiac conduction disease; CIED = cardiac implantable electronic device; CMP = cardiomyopathy; CV = cardiovascular; DCM = dilated cardiomyopathy; FHx = family history; HCM = hypertrophic cardiomyopathy; LVAD = left ventricular assist device; RCM = restrictive cardiomyopathy; SCD = sudden cardiac death; VT = ventricular tachycardia; VF = ventricular fibrillation.

### Genetic Landscape

Missense variants were identified in 203 (88.3%), and non-missense variants in 27 (11.7%) patients (see **Table S3** for list of variants included in the study). Notably, 12 (5.2%) carried homozygous variants (7 non-missense and 5 missense) and 5 (2.2%) hosted compound heterozygous *DES* variants (3 patients had 2 different non-missense alleles, and 2 had a non-missense as well as a missense variant). Fifteen additional patients carried 9 unique non-missense variants in heterozygous state. Nine (3.9%) patients carried confirmed *de novo* variants. The variant locations across *DES* from the patients included in the study as well as associated pleiotropic cardiomyopathy and skeletal myopathy phenotypes are shown in **Figure 1**. Patients with variants in the 2B domain were the most frequent (n=79, 34.3%), followed by patients with head or tail domains variants (n=42, 18.3% each).

**Figure 1.**
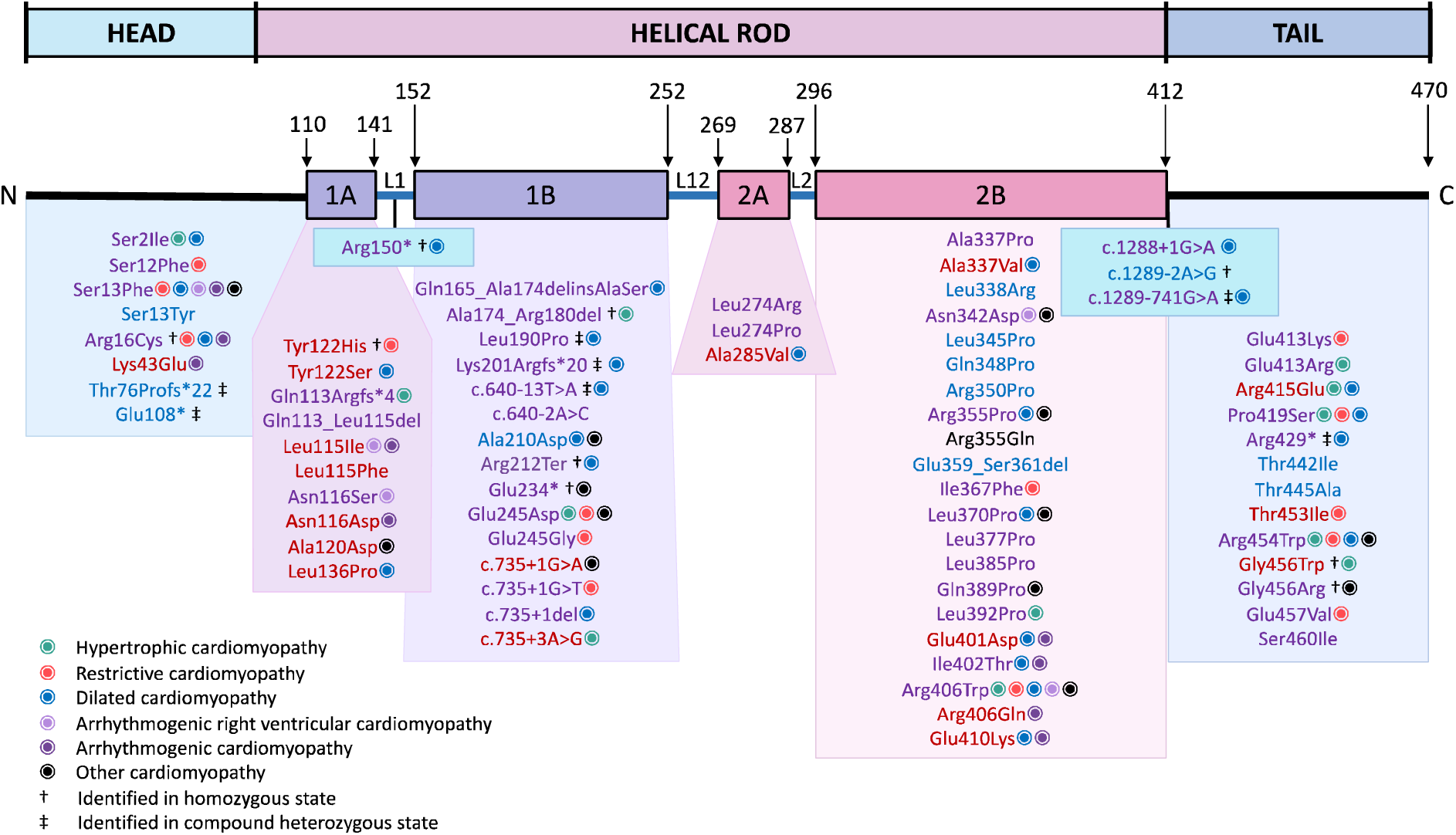
**Structural organization of the human desmin (DES) gene and localization of variants included in this study.** Pathogenic variants are found across the entire *DES* gene, with majority of patients carrying variants localized in the α-helical-rod 2B and in the tail domains. Variants are classified into three groups depending on the ensuing phenotype: cardiac involvement (**red**), skeletal myopathy (**blue**), both cardiac involvement and skeletal myopathy (**purple**), and neither (**black**). Associated cardiomyopathy phenotypes are provided with additional markers (see figure legend). † Considered carrier status in heterozygous state, but pathogenic or likely pathogenic in homozygous state. ‡ Considered carrier status unless identified in compound heterozygous state. 1A and 1B, coil 1A and coil 1B; 2A and 2B, coil 2A and coil 2B; L1, L12 and L2, non-alpha-helical linker regions.

### Phenotype Spectrum

Out of 230 patients, 89 (38.7%) had cardiac and skeletal muscle involvement, an additional 79 (34.3%) patients showed only cardiac involvement, 39 (17.0%) had only skeletal muscle involvement, and 23 (10.0%) had neither phenotype. Overall, 124 (53.9%) patients were diagnosed with a cardiomyopathy either at baseline or during follow-up. Dilated cardiomyopathy (DCM) was the most common form of cardiomyopathy (14.8% of all patients), followed by restrictive cardiomyopathy (RCM, 13.5% of all patients), whereas the remaining forms were less common: arrhythmogenic cardiomyopathy (ACM, 7.0% of all patients), hypertrophic cardiomyopathy (HCM,6.1% of all patients), arrhythmogenic right ventricular cardiomyopathy (ARVC, 5.2% of all patients), and other forms (7.4% of all patients). Twenty-seven (11.7%) patients developed atrial fibrillation at median age of 43.0 years [24.0; 53.5].

Out of 134 patients who had no cardiomyopathy phenotype at baseline evaluation, 35 (26.1%) presented with CCD (median age at presentation 32.0 years [23.0; 40.0]. Of them, 13/35 (37.1%) were diagnosed with a cardiomyopathy during follow-up: 6 developed RCM, 5 progressed to DCM, and 2 developed other forms of cardiomyopathies.

Overall, 76 patients (33.0%) had only a single clinical encounter, and follow-up information was available in 154 (67.0%). Median follow-up time was 3 (0; 11.0) years. Transition of cardiomyopathy phenotype during follow-up was seen in 11/76 (14.5%) patients (6 with other cardiomyopathy [cmp] and 1 patient with HCM had phenotype transition to DCM, and 2 patients with HCM and 1 with other CMP had transition to RCM), including 2 patients with first presentation in childhood. Median time from initial presentation to transition of cardiomyopathy phenotype was 10 years (3.5; 18.5).

### Familial Cardiac Disease Penetrance

Probands were more likely to have non-missense *DES* variants (17.5% vs 5.5%, p=0.005). Out of 21 probands with non-missense *DES* variants, 10 (47.6%) were found to be in homozygous or compound heterozygous states with another *DES* variant. In contrast, only 15 (12.5%) out of 120 probands with missense *DES* variants had a homozygous genotype or co-occurred with another *DES* variant. Probands more frequently had cardiac involvement (defined as cardiomyopathy phenotype and/or MACE) than family members (81.7% vs 63.6%, p=0.002), whereas no significant difference was observed between males and females (77.7% vs 67.9%, p=0.103).

### Major Adverse Cardiac Events (MACE): CCD, Sustained VA and HF Events

Overall, 132 (57.4%) patients with P/LP *DES* variants experienced at least one MACE event, with an event rate per person-year of 1.59% [1.32%–1.87%] (Figure 2). In 83 (36.1%) patients, MACE was the first manifestation of disease. Notably, 37 (16.1%) patients experienced more than one type of outcome. The median age at first MACE was 36.5 [24.0; 48.0] years, with probands being, on average, 8 years younger than relatives at the time of their first MACE event (30.0 vs. 38.0 years, p=0.012). No sex-related differences were observed in the risk or age at composite MACE (Figure 3).

**Figure 2.**
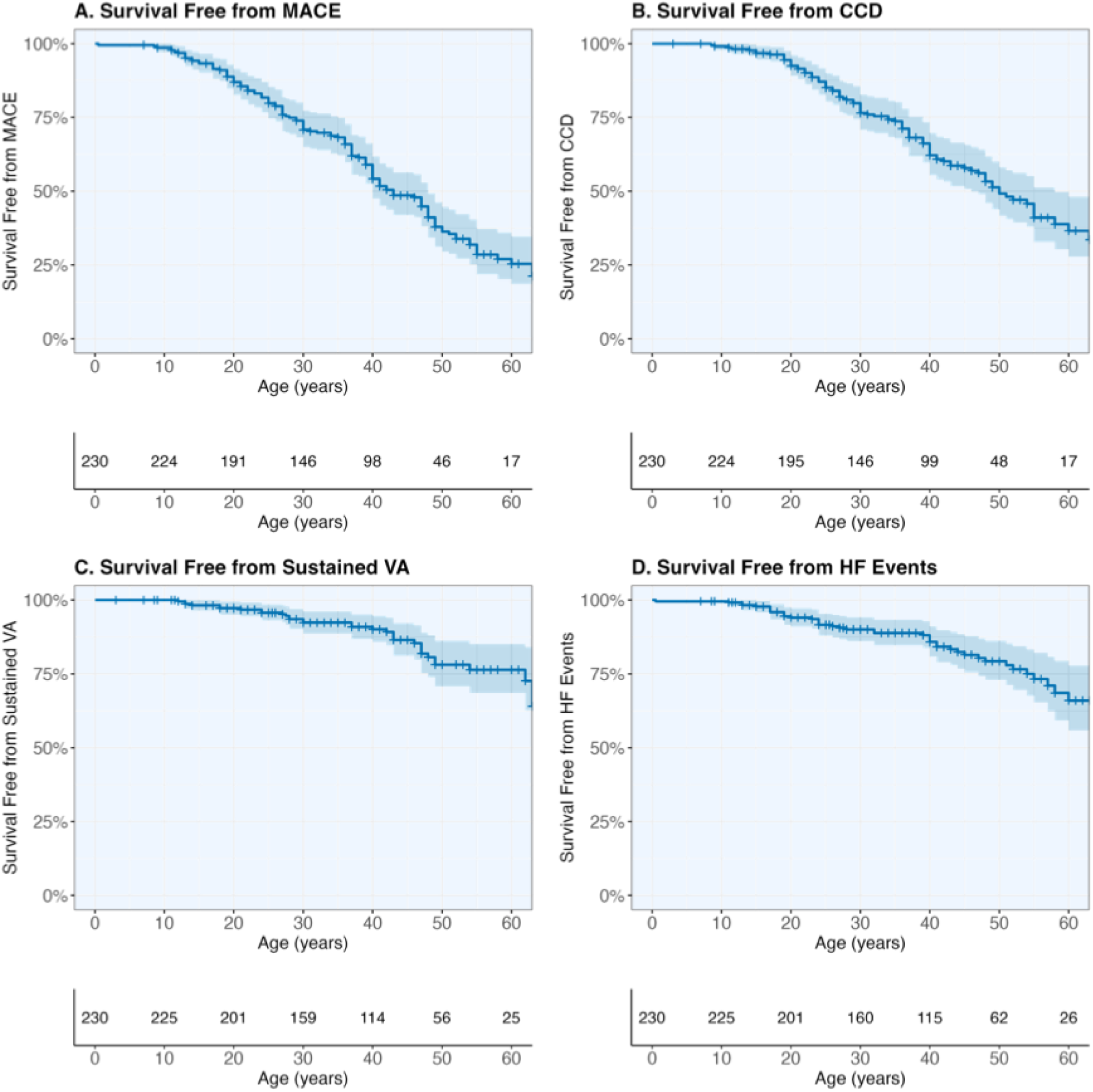
**Survival free from major adverse cardiac events (MACE).** Survival free from composite MACE (**A**), cardiac conduction disease (CCD) requiring cardiac implantable electronic device (CIED) implantation (**B**), sustained ventricular arrhythmia (VA) events, (**C**), and heart failure (HF) events (HF hospitalization, left ventricular assist device (LVAD)/cardiac transplantation, HF-related death) (**D**) are presented with 95% CIs (shaded area) in the overall population.

**Figure 3.**
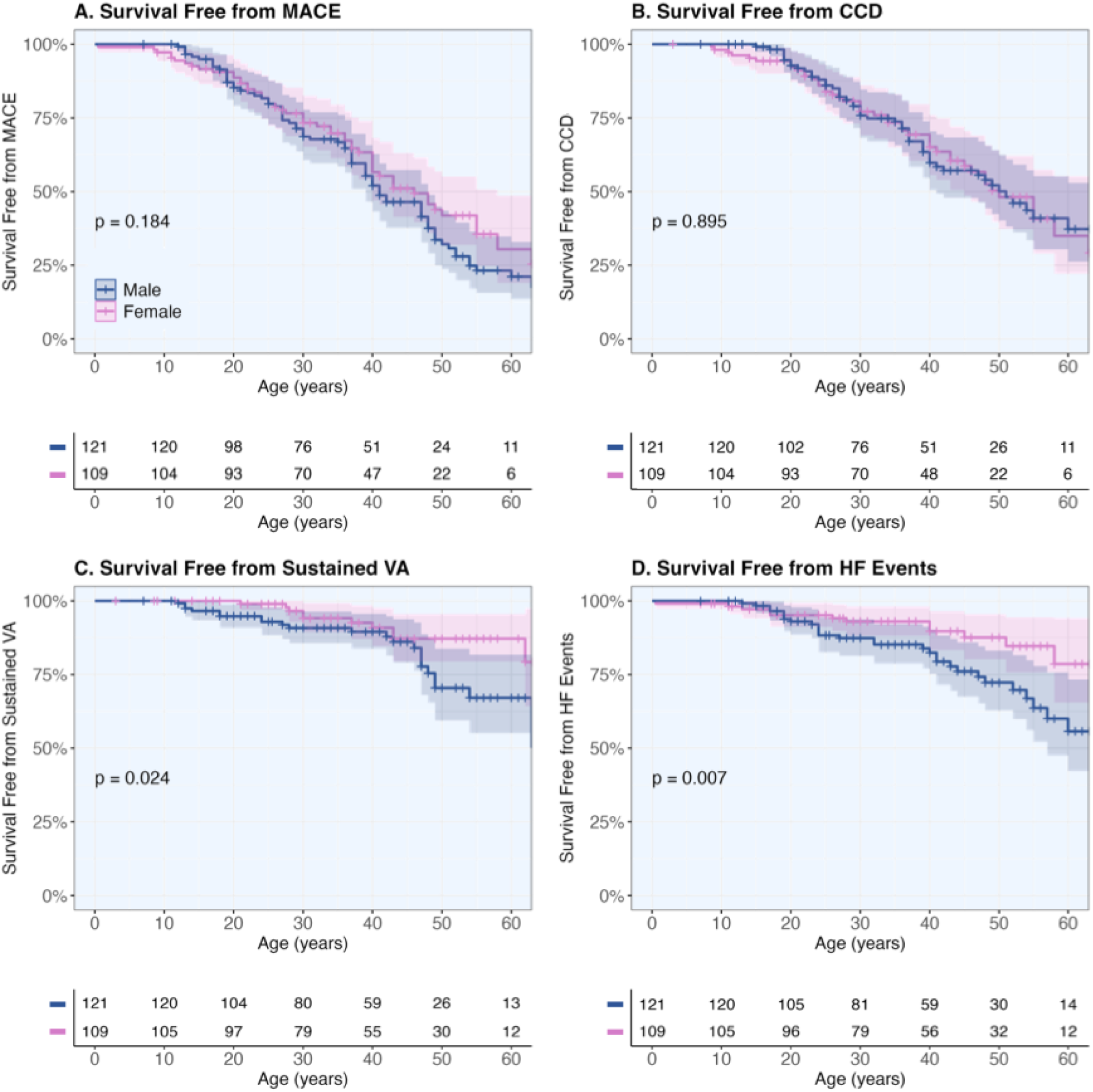
**Survival free from major adverse cardiac events (MACE) stratified by sex.** Survival free from composite MACE (**A**), cardiac conduction disease (CCD) requiring cardiac implantable electronic device implantation (**B**), sustained ventricular arrhythmia (VA) events, (**C**), and heart failure (HF) events (HF hospitalization, left ventricular assist device (LVAD)/cardiac transplantation, HF-related death) (**D**) are presented with 95% CIs (shaded area) in males vs females.

In our study, the most common outcome was CCD (n=96/230, 41.7%), followed by HF events (n=43/230, 18.7%) and sustained VA events (n=36/230, 15.7%) (Table 1). Among 96 patients who had CCD, 66 (68.8%) had complete AV block, 19 (19.8%) developed an unspecified degree of AV block requiring CIED implantation, and 5 (5.2%) had advanced AV block; the remaining 6 patients had other conduction disturbances, which required CIED implantation. Among 36 patients who had sustained VA events, 19 (52.8%) had sustained VT, 4 (11.1%) survived after VF, 8 (22.2%) had appropriate ICD therapy, and 5 (13.9%) succumbed to SCD. Among 43 patients who had HF events, 23 (53.5%) had HF hospitalization, 19 (44.2%) underwent LVAD implant or cardiac transplant for advanced HF, and 13 (30.2%) had HF-related death. In univariable Cox proportional hazards analyses, male sex was associated with increased risk for sustained VA (HR: 2.28; [95%CI: 1.12–4.64]) and HF events (HR: 2.45; [95%CI: 1.26–4.77]) but not for CCD (HR: 0.97; [95%CI: 0.65–1.46] or MACE (HR: 1.27; [95%CI: 0.89–1.79].

Analysis of outcome rates per cardiomyopathy phenotype revealed significant differences (Figure 4). Patients with ACM phenotype were more likely and those with HCM phenotype less likely to develop sustained VA events. On the other hand, CCD events were most frequently observed in those with DCM or RCM phenotype, less common among those with ARVC, and not represented in those with ACM phenotype.

**Figure 4.**
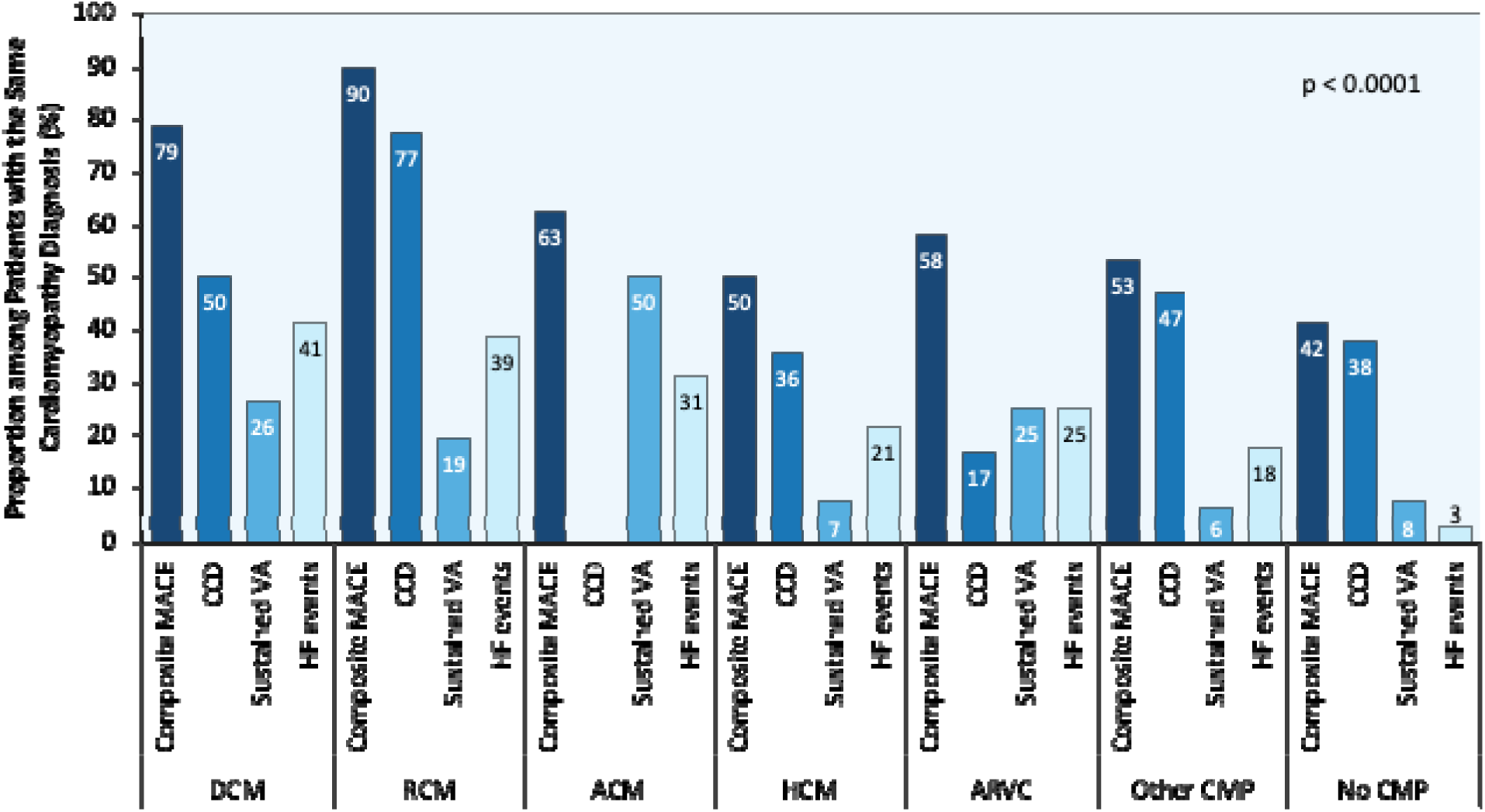
**Proportion of patients with individual and composite major adverse cardiac events (MACE) stratified by the underlying cardiomyopathy phenotype.** Note, for patients with transition of phenotype, the more advanced phenotype stage was chosen for this analysis. ACM = arrhythmogenic cardiomyopathy; ARVC = arrhythmogenic right ventricular cardiomyopathy; CMP = cardiomyopathy; DCM = dilated cardiomyopathy; HCM = hypertrophic cardiomyopathy; RCM = restrictive cardiomyopathy.

Overall, 26 (11.3%) patients died during follow-up, including 19 (8.3%) patients who died of a cardiovascular cause. Median age at death was 40.0 years (28.0; 52.0) (**Figure S2**).

## DISCUSSION

The growing focus on genotype-centered diagnosis and management of cardiomyopathies has exposed a critical gap in our understanding of *DES*-associated cardiomyopathy and highlights the need for rigorous and comprehensive research to define the care of patients affected with this disease.(13,14) Efforts to date have been impeded by the rarity of *DES*-associated cardiomyopathy and its varied manifestations, which often lead patients to seek care across multiple clinical disciplines. To address this critical need, we conducted a systematic review and literature-based individual patient data meta-analysis of individuals harboring P/LP *DES* variants to comprehensively describe the genetic landscape, natural history, phenotype spectrum, familial disease penetrance and outcomes of *DES*-associated cardiomyopathy. Our large-scale analysis, encompassing data from 230 patients with 74 unique *DES* variants from 71 publications, has yielded significant insights that hold potential to reshape clinical care within this population. More than half (53.9%) of patients with P/LP *DES* variants exhibited a cardiomyopathy phenotype, often accompanied by features of skeletal myopathy. We observed marked heterogeneity of cardiomyopathy phenotypes, with DCM emerging as the predominant form, followed by RCM, while other phenotypes were less common. Of profound concern, 57.4% of patients with P/LP *DES* variants experienced MACE events, with CCD requiring CIED implantation being the most common (41.7%), followed by HF events (18.7%) and sustained VA events (15.6%). The median age at MACE was 36.5 years, underscoring the profound impact of *DES*-associated cardiomyopathy on morbidity and mortality at relatively young ages. Furthermore, we observed incomplete yet high familial cardiac penetrance of cardiac disease, with nearly two-thirds of relatives harboring a P/LP *DES* variant developing cardiomyopathy or experiencing MACE events. Finally, sex emerged as a modifier of outcomes, with male patients showing heightened susceptibility to sustained VA and HF events. These factors should, therefore, be integral to risk stratification and management strategies in patients with *DES* variants.

These insights redefine *DES*-associated cardiomyopathy as a distinct rare disease typified by early onset, diverse cardiomyopathy phenotypes often accompanied by skeletal myopathy, high familial penetrance, and a significant burden of MACE (**SUMMARY FIGURE**).

### *DES* Variants Cause Heterogeneous Cardiomyopathy Phenotypes

Since the initial identification of pathogenic *DES* variants in 1998,(20,23–25) multiple *DES* variants across the gene have been reported. P/LP *DES* variants typically disrupt desmin filament assembly at different steps.(26,27) This disruption leads to intra-cellular deposition of osmiophilic granulofilamentous material that is immunoreactive with anti-desmin antibodies in both myocardial and skeletal muscle tissues.(28) Studies conducted with genetically modified cell lines with human *DES* variants or in *DES* knockout mice have demonstrated that desmin critically modulates mitochondrial functions.(29) Consequently, mutated desmin has been shown to lead to severe mitochondrial abnormalities that directly contribute to the development of *DES-*associated (cardio)myopathy. Early studies indicated that *DES* variants are causally implicated in approximately 2% of all DCM cases.(15) However, given the marked heterogeneity of *DES*-associated phenotypes, this proportion likely underestimates disease prevalence in cohorts with diverse cardiomyopathy phenotypes, necessitating more accurate estimates.

To our knowledge, this study represents the first large-scale description of the natural history of *DES-*associated cardiac disease, providing data on familial penetrance and outcomes. Our findings reveal marked phenotype heterogeneity in *DES*-associated cardiomyopathy, predominantly presenting as DCM or RCM, with ≥80% of these cases developing advanced or complete AV block. Conversely, ACM, HCM and ARVC phenotypes were less common. Of note, disease expression varied not only across different genetic variants but also within and between families carrying the same variant. A recent study investigated the phenotype and clinical outcomes in patients with *DES* variants, including only individuals meeting the Padua criteria for ACM.(30) As a result of the selective phenotype inclusion, this cohort lacked patients with HCM/RCM phenotypes or CCD. There was nearly complete absence of myopathy phenotype in this population, further indicating the incomplete depiction of *DES-*associated disease.(4) Our findings suggest that the phenotype defined by any single existing cardiomyopathy diagnostic scheme would capture only a portion of the breadth of *DES*-associated cardiac disease expressions. Using limited longitudinal follow-up data, we further demonstrate that transition of cardiomyopathy phenotype in patients with *DES*-associated cardiac disease is uncommon. Prior reports suggest that phenotype transition may occur in some children with HCM progressing to RCM (often with CCD) and evolving towards a dilated phase at advanced stages.(31) Notably, while DCM with associated AV conduction disease is commonly observed in other syndromic cardiomyopathies with neuromuscular manifestations (e.g. *LMNA* and *EMD* disease),(7) the combination of RCM with advanced/complete AV block is highly indicative of *DES-*associated disease.(28) Once cardiac amyloidosis as the most frequent cause of RCM is excluded by appropriate diagnostic work-up, the combination of RCM and advanced/complete AVB strongly suggests *DES*-associated cardiomyopathy.(25,28) Skeletal myopathy, whether clinically manifest or subclinical (identified by biopsy), when associated with RCM and/or AV block should also raise suspicion of *DES*-associated disease. Notably, the skeletal myopathy associated with *DES* variants is heterogeneous, as it can present with either distal or proximal weakness, making it harder to recognize.

### *DES* Variants Demonstrate a High Familial Penetrance

To date, the evaluation of familial penetrance of *DES*-associated cardiomyopathy has been limited to individual families or founder populations,(32) and larger-scale analyses have been lacking. In this study, the disease inheritance appeared autosomal dominant in the vast majority of cases, and autosomal recessive or sporadic in a minority of cases, consistent with prior studies.(32–34) Our data indicate a substantial familial disease burden, with 20% of probands with *DES* variants having a family history of cardiomyopathy, one in eight having a relative with SCD, and one-fourth having a relative with a CIED. Moreover, P/LP *DES* variants caused a highly penetrant cardiac disease, with nearly two-thirds either being diagnosed clinically with cardiomyopathy or manifesting a MACE. This familial clustering and high penetrance underscore the importance of timely genetic evaluation in affected families to enable pre- clinical identification of asymptomatic at-risk relatives. However, longitudinal studies are necessary regarding the predictors and probability of development of cardiomyopathy or MACE over time among at-risk relatives before specific recommendations can be developed.(35)

### *DES*-Associated Cardiomyopathy is Associated with High Burden of MACE

Nearly 60% of patients in our cohort experienced at least one MACE event, most frequently advanced/complete AV block requiring CIED implantation. The median age of 36.5 years at MACE indicates a high burden of potentially lethal events at a relatively young age. Of note, 36% of all patients presented with MACE as the first manifestation of disease, underscoring the potential benefits of cascade screening and early diagnosis.

Male patients with *DES* variants were at increased risk of both sustained VA and HF events, as compared to females. The association of male sex with higher risk of sustained VA is also recognized in several other cardiomyopathies, including *LMNA, EMD, PKP2* and *TMEM43* disease,(36–39) and has been recently proposed for patients with *DES* variants and ACM phenotype.(30) This is in contrast to desmoplakin (*DSP*) cardiomyopathy where female sex was recently demonstrated to be associated with higher risk of sustained VA events.(40) Mechanisms responsible for the higher risk of sustained VA and HF events in male patients with *DES* variants have not been explored.(15)

When analyzing individual outcomes, we found that over one-fourth of subjects presenting with unexplained CCD requiring CIED implantation were diagnosed with a cardiomyopathy at follow-up. Mechanisms underlying the early development of CCD before cardiomyopathy manifests remain insufficiently studied.(41) It has been proposed that *DES*-associated AV block may be caused by anatomical interruption of the AV conduction system related to fibrosis or calcification. A post-mortem autopsy of one Japanese patient with the p.A337P variant showed calcification of the His bundle and sporadic calcium deposits in the left and right bundle branches.(42) Conversely, another autopsy case of a young adult male, who was diagnosed with complete AV block at the age of 13 and died from advanced RCM in his 20s, showed no calcification of the compact AV node or the His bundle. Instead, it revealed extensive fibrosis of the terminal portions of the branching bundle and the top of the ventricular septum, corresponding to the initial segments of the left and right bundles.(43) As desmin has been shown to critically modulate mitochondrial functions,(29) the early development of CCD may also be the consequence of mitochondrial abnormalities in Purkinje fibers caused by mutant desmin protein. The consequent development of cardiomyopathy in *DES* patients with CCD illustrates the progressive nature of the disease and emphasizes the necessity for comprehensive clinical and genetic evaluations in young and middle-aged patients presenting with unexplained CCD to facilitate the timely diagnosis and effective management of patients with underlying genetic diseases. This need has been recently acknowledged by the international consensus statement, with incorporation of new recommendations for genetic testing in CCD.(44) However, *DES* as a cause of isolated CCD or CCD combined with non-DCM phenotypes remains underrecognized.(44) Therefore, clinicians should ensure that *DES* is included in the panel of genes tested during genetic evaluations.

Our findings suggest that male patients with *DES* variants in particular should have a low threshold for primary preventive ICD implantation. Presence of ARVC/ACM phenotype should further support this notion. Additionally, considering the knowledge in the field form other cardiomyopathies, the clinical presentation and history might contribute to decision making in both sexes, with history of arrhythmic syncope, high burden of late gadolinium enhancement on CMR, or family history of SCD possibly being considered likely surrogates of VA risk, before evidence-based recommendations are available. In view of the phenotypic overlap between *DES*- and *LMNA*-associated cardiomyopathies, such as early development of CCD and high burden of sustained VA and HF events, it is notable that male sex is also included in the *LMNA* risk calculator for primary prevention,(37) further underscoring the similarities between the two diseases. Although the potential association between CCD and the risk of sustained VA events in *DES*-associated cardiomyopathy remains unexplored, we recommend considering primary prevention ICD implantation for patients with CCD who require a pacemaker.

### Clinical Implications

Our findings indicate that both probands and family members with P/LP *DES* exhibit a severe phenotype, characterized by a high burden of MACE and high mortality at a relatively young age primarily due to cardiac complications. These events include CCD typically early in the disease, sustained VA and advanced HF events. We therefore emphasize the importance of comprehensive genetic evaluation in families suspected of having *DES* disease, including those with otherwise unexplained CCD-PM at a young and middle age, to inform clinical decision making. Male patients are at an increased risk for sustained VA events, and this association should be considered when making clinical decisions regarding primary preventive ICD implantation. Since patients with P/LP *DES* variants often experience multiple and diverse MACE, early involvement of relevant subspecialty care domains is advisable to ensure these patients receive best-practice multidisciplinary and precision care.

### Future Directions

To advance the understanding and management of *DES*-associated cardiomyopathy, several key areas require focused research. Whereas in other cardiomyopathies, such as *FLNC*(45) and *MYH7* variant type determines the ensuing cardiomyopathy phenotype,(34,46,47) we found that select *DES* variants demonstrate high pleiotropy of cardiac phenotypes. We also found that non-missense variants were 4 times more likely to be found in the context of homozygous or compound heterozygous states, than missense variants. This calls into question the lower pathogenicity of the non-missense variants. To this end, whether loss of function is a mechanism of DES-associated disease, remains to be established. More research is necessary to elucidate the mechanisms governing pleiotropic phenotypes and sex-related differences in disease expression and outcomes. Longitudinal studies are essential to monitor at-risk relatives over time, assessing the factors that govern the development of cardiomyopathy and MACE to inform early intervention strategies. Future research should also include diverse populations to understand the role of ethnicity in disease presentation and outcomes. Collaborative efforts to establish multicenter registries and incorporate biobank-level insights into causal genetic architecture of disease will enhance the understanding of genotype-phenotype correlations and promote personalized medicine approaches. Finally, improving clinical awareness about *DES*-associated cardiomyopathy is vital to mitigating disease progression and improving patient outcomes.

### Study Strengths and Limitations

To our knowledge, this is the largest-scale study of patients with P/LP *DES* variants to date. However, our findings should be viewed in the light of its systematic evidence-based review design and heterogeneous reporting of included studies as limitations that affected the quality of evidence in our analysis. As such, our reliance on reported phenotypes and outcomes restricted the description of skeletal myopathy and *DES*-associated cardiomyopathy phenotypes. The study design also hindered analysis of prescribed medications and guideline-directed medical therapy; however, to date, no medication has been reported to alter disease outcomes in *DES*-associated cardiomyopathy.

Our analysis included both founder and non-founder variants. Additionally, the majority of patients included in our study lacked reported ancestry information, precluding an assessment of the role of ethnicity in phenotype and outcomes. Current ACMG/AMP recommendations used to classify rare variants in *DES* required excluding some VUS reported in association with cardiac and/or skeletal myopathy phenotypes in the literature (albeit with no evidence of segregation with phenotype). This is particularly relevant for heterozygous non-missense variants as currently loss of function as a mechanism of disease is not unequivocally established. Therefore, our cohort does not account for the phenotype observed in cases with VUS; some of these VUS may be upgraded to P/LP classification upon the availability of more variant-level data or gene–disease association evidence.

Furthermore, because we included only patients who presented alive, the outcome exposure, particularly regarding sustained VA and mortality, may potentially underestimate real-world outcomes in patients with P/LP *DES* variants. The phenotypic differences between probands and relatives may also be partly explained by publication bias. Similarly, as familial penetrance was calculated based on patients who underwent clinical cardiac evaluation, an ascertainment bias cannot be excluded. Finally, as our findings are derived from patients with P/LP *DES* variants diagnosed in the clinical setting, the ensuing implications should not be directly extrapolated to individuals with P/LP *DES* identified as incidental or secondary findings, as the disease penetrance and outcomes in this population remain poorly investigated.(48)

## CONCLUSIONS

This study establishes *DES* cardiomyopathy as a distinct genetic heart disease characterized by heterogeneous phenotypes, often accompanied by skeletal myopathy, and significant MACE burden, including CCD (often preceding cardiomyopathy phenotype), sustained VA and advanced HF events. The high familial penetrance and early onset of severe events emphasize the need for cascade screening and close monitoring in affected families. Male sex was identified as a risk modifier and can be used to guide risk stratification and management strategies. Our findings highlight the necessity for individualized management approaches in managing *DES* cardiomyopathy, incorporating genetic information and genotype-specific risk factors to optimize patient outcomes. Future studies should focus on elucidating the mechanisms driving disease expression and developing genotype-tailored risk models that capture the natural history of *DES* cardiomyopathy.

## Data Availability

A limited dataset will be made available upon reasonable request.

## Supplementary Material

Tables S1-S3 Figure S1-S2

## Nonstandard Abbreviations And Acronyms

ACM: arrhythmogenic cardiomyopathy
ACMG: American College of Medical Genetics and Genomics
AMP: Association for Molecular Pathology
P/LP: pathogenic / likely pathogenic
VA: ventricular arrhythmia
VUS: variant(s) of uncertain significance

---

**Summary Figure.**
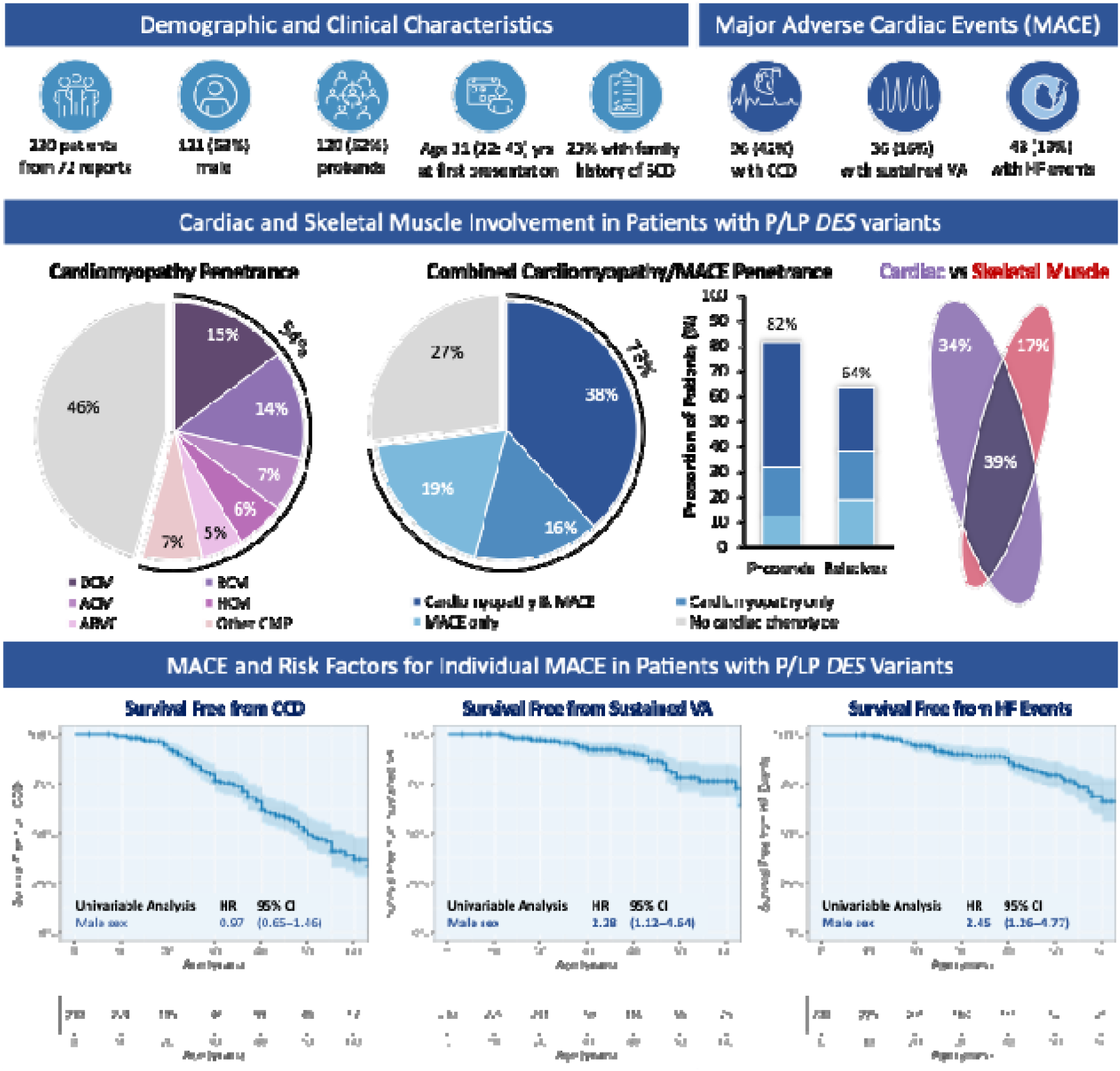
The Distinct Natural History, Phenotype Spectrum, Familial Penetrance and Clinical Outcomes in Desmin (*DES*)-Associated Cardiomyopathy.

