## Supplementary material for "Natural History, Phenotype Spectrum and Clinical Outcomes of Desmin (*DES*)-Associated Cardiomyopathy": Table S1

**SUPPLEMENTARY MATERIAL**

### Table S1. Search strategy in databases.

| **A. PubMed Search, 1998 to present, English** | |
| --- | --- |
| ("Desmin"[MeSH Terms] OR "Desmin"[Tiab] OR "DES"[Tiab] NOT ("diethylstilbestrol"[tiab])) AND ("Mutation"[MeSH Terms] OR "DNA Mutational Analysis"[MeSH Terms] OR "pathogenic variant*"[Tiab] OR "pathogenic"[Tiab] OR "mutat*"[Tiab] OR "desminopath*"[Tiab] OR "variant*"[Tiab] OR "defect*"[Tiab] OR "myopath*"[Tiab] OR "deletion*"[Tiab]) NOT ("animals"[MeSH] NOT ("animals"[MeSH] AND "humans"[MeSH])) AND (english[Filter]) AND (1998:2024[pdat]) | |
| **Results:** 2156 results. **Date:** 5/31/2024 | |
| **B. Embase, 1998 to present** | |
| (('desmin'/exp OR 'desmin':ti,ab,kw OR 'des':ti,ab,kw) NOT 'diethylstilbestrol':ti,ab,kw) AND ('mutation'/exp OR 'dna mutational analysis'/exp OR 'pathogenic variant*':ti,ab,kw OR 'pathogenic':ti,ab,kw OR 'mutat*':ti,ab,kw OR 'desminopath*':ti,ab,kw OR 'variant*':ti,ab,kw OR 'defect*':ti,ab,kw OR 'myopath*':ti,ab,kw OR 'deletion*':ti,ab,kw) NOT (('animal'/exp OR 'animal experiment'/exp OR 'nonhuman'/exp) NOT ('human'/exp OR 'human experiment'/exp)) AND [english]/lim AND [1998-2024]/py | |
| **Results:** 3590 results. **Date:** 5/31/2024 | |
| **Search Summary** | **Citations** |
| PubMed | 2,156 |
| Embase | 3,590 |
| **Sub-Total** | **5,746** |
| Duplicates removed (Endnote) | 1,534 |
| Duplicates removed (Covidence) | 39 |
| **Total citations** | **4,212** |

### Table S2. List of publications included in the systematic review and individual patient data meta-analysis.

| Study ID | Author, year | Year | N of probands included | N of relatives included |
| --- | --- | --- | --- | --- |
| 1. | Muñoz-Mármol et al.(1) | 1998 | 1 | – |
| 2. | Sjöberg et al.(2) | 1999 | 1 | – |
| 3. | Sugawara et al.(3) | 2000 | 1 | – |
| 4. | Goudeau et al.(4) | 2001 | 1 | – |
| 5. | Dagvadorj et al.(5) | 2004 | 2 | – |
| 6. | Vrabie et al.(6) | 2005 | 1 | – |
| 7. | Bär et al.(7) | 2005 | 1 | – |
| 8. | Fidziańska et al.(8) | 2005 | 1 | – |
| 9. | Arbustini et al.(9) | 2006 | 3 | 1 |
| 10. | Hager et al.(10) | 2006 | 1 | – |
| 11. | Arias et al.(11) | 2006 | 1 | 2 |
| 12. | Pruszczyk et al.(12) | 2007 | 1 | 2 |
| 13. | Yuri et al.(13) | 2007 | 1 | – |
| 14. | Bär et al.(14) | 2007 | 1 | – |
| 15. | Olivé et al.(15) | 2007 | 3 | 1 |
| 16. | Strach et al.(16) | 2008 | 4 | 8 |
| 17. | Piñol-Ripoll et al.(17) | 2009 | 1 | – |
| 18. | van Tintelen et al.(18) | 2009 | 5 | 14 |
| 19. | Otten et al.(19) | 2010 | 1 | 1 |
| 20. | Hong et al.(20) | 2011 | 7 | 15 |
| 21. | Klauke et al.(21) | 2010 | 1 | – |
| 22. | He et al.(22) | 2010 | 1 | – |
| 23. | Wahbi et al.(23) | 2012 | 14 | 7 |
| 24. | van Spaendonck-Zwarts et al.(24) | 2012 | 6 | 9 |
| 25. | Tse et al.(25) | 2013 | 1 | – |
| 26. | Henderson et al.(26) | 2013 | 1 | – |
| 27. | Cetin et al.(27) | 2013 | 1 | – |
| 28. | McLaughlin et al.(28) | 2013 | 1 | – |
| 29. | Palmio et al.(29) | 2013 | 1 | – |
| 30. | Brodehl et al.(30) | 2013 | 1 | – |
| 31. | Nalini et al.(31) | 2013 | 1 | 3 |
| 32. | Fichna et al.(32) | 2014 | 0 | 1 |
| 33. | Ripoll-Vera et al.(33) | 2015 | 3 | 11 |
| 34. | Faksh et al.(34) | 2015 | 1 | – |
| 35. | Brodehl et al.(35) | 2016 | 1 | – |
| 36. | Boulé et al.(36) | 2015 | 1 | – |
| 37. | Ojrzyńska et al.(37) | 2017 | 1 | – |
| 38. | Bermúdez-Jiménez et al.(38) | 2018 | 1 | 20 |
| 39. | Schirmer et al.(39) | 2018 | 1 | – |
| 40. | Gearhart et al.(40) | 2018 | 1 | – |
| 41. | Oomen et al.(41) | 2018 | 2 | – |
| 42. | Fan et al.(42) | 2019 | 1 | – |
| 43. | Marakhonov et al.(43) | 2019 | 1 | 1 |
| 44. | Riley et al.(44) | 2019 | 1 | 1 |
| 45. | Stępień-Wojno et al.(45) | 2020 | 1 | – |
| 46. | Brodehl et al.(46) | 2019 | 1 | – |
| 47. | Hsu et al.(47) | 2019 | 1 | 1 |
| 48. | Nicolau et al.(48) | 2019 | 1 | – |
| 49. | Tamiya et al.(49) | 2020 | 1 | – |
| 50. | Kubánek et al.(50) | 2020 | 4 | 1 |
| 51. | Protonotarios et al.(51) | 2021 | 2 | 6 |
| 52. | Fischer et al.(52) | 2021 | 2 | 2 |
| 53. | Chen et al.(53) | 2021 | 1 | – |
| 54. | Potulska-Chromik et al.(54) | 2021 | 1 | – |
| 55. | Huang et al.(55) | 2021 | 1 | – |
| 56. | Santhoshkumar et al.(56) | 2021 | 1 | – |
| 57. | Chen et al.(57) | 2021 | 1 | – |
| 58. | Brodehl et al.(58) | 2021 | 1 | – |
| 59. | Oka et al.(59) | 2021 | 1 | – |
| 60. | Claes et al.(60) | 2023 | 1 | 1 |
| 61. | Rauf et al.(61) | 2022 | 1 | – |
| 62. | Liu et al.(62) | 2022 | 1 | – |
| 63. | Onore et al.(63) | 2022 | 1 | – |
| 64. | Yeow et al.(64) | 2022 | 2 | 1 |
| 65. | Takegami et al.(65) | 2023 | 1 | – |
| 66. | Dias et al.(66) | 2023 | 1 | – |
| 67. | Monda et al.(67) | 2023 | 1 | – |
| 68. | Papadopoulos et al.(68) | 2023 | 1 | – |
| 69. | Geist Hauserman et al.(69) | 2024 | 1 | – |
| 70. | Bučić et al.(70) | 2024 | 1 | – |
| 71. | Bermudez-Jimenez et al.(71) | 2024 | 6 | 1 |

### Table S3. List of *DES* variants included in the study.^*^

| Variant ID | Nucleotide change | Protein change | dbSNP ID | N of patients | Genotype (Het/CH/Hom) |
| --- | --- | --- | --- | --- | --- |
| 1. | c.5G>T | p.Ser2Ile | rs58999456 | 3 | heterozygous |
| 2. | c.35C>T | p.Ser12Phe | rs267607495 | 4 | heterozygous |
| 3. | c.38C>T | p.Ser13Phe | rs62636495 | 29 | heterozygous |
| 4. | c.38C>A | p.Ser13Tyr | rs62636495 | 1 | heterozygous |
| 5. | c.46C>T | p.Arg16Cys | rs60798368 | 3 | homozygous |
| 6. | c.127A>G | p.Lys43Glu | rs397516689 | 1 | heterozygous |
| 7.^**^ | c.226del | p.Thr76Profs*22 | rs1399282762 | 1 | compound heterozygous |
| 8.^**^ | c.322G>T | p.Glu108* | rs62636490 | 1 | compound heterozygous |
| 9. | c.338_339del | p.Gln113Argfs*4 | rs267607497 | 2 | heterozygous |
| 10. | c.336_344del | p.Gln113_Leu115del | rs1553603239 | 2 | heterozygous |
| 11. | c.343C>A | p.Leu115Ile | n.a. | 10 | heterozygous |
| 12. | c.343C>T | p.Leu115Phe | n.a. | 2 | heterozygous |
| 13. | c.347A>G | p.Asn116Ser | rs267607499 | 1 | heterozygous |
| 14. | c.346A>G | p.Asn116Asp | n.a. | 1 | heterozygous |
| 15. | c.359C>A | p.Ala120Asp | rs1954373010 | 1 | heterozygous |
| 16. | c.364T>C | p.Tyr122His | n.a. | 1 | homozygous |
| 17. | c.365A>C | p.Tyr122Ser | n.a. | 1 | heterozygous |
| 18. | c.407T>C | p.Leu136Pro | rs397516695 | 1 | heterozygous |
| 19.^**^ | c.448C>T | p.Arg150* | n.a. | 1 | homozygous |
| 20. | c.493_520delinsGCGT | p.Gln165_Ala174delinsAlaSer | rs1114167332 | 1 | heterozygous |
| 21.^**^ | c.521_541del | p.Ala174_Arg180del | rs60538473 | 2 | homozygous |
| 22.^**^ | c.569T>C | p.Leu190Pro | rs1243057653 | 2 | compound heterozygous |
| 23.^**^ | c.600del | p.Lys201Argfs*20 | rs727504448 | 1 | compound heterozygous |
| 24. | c.640-2A>C |  | rs267607492 | 2 | heterozygous |
| 25. | c.629C>A | p.Ala210Asp | n.a. | 2 | heterozygous |
| 26.^**^ | c.634C>T | p.Arg212* | rs781590560 | 1 | homozygous |
| 27.^**^ | c.640-13T>A |  |  | 1 | compound heterozygous |
| 28.^**^ | c.700G>T | p.Glu234* | n.a. | 2 | homozygous |
| 29. | c.735G>C | p.Glu245Asp | rs267607486 | 10 | heterozygous |
| 30. | c.734A>G | p.Glu245Gly | rs1575014243 | 5 | heterozygous |
| 31. | c.735+1G>A |  | rs397516698 | 1 | heterozygous |
| 32. | c.1024A>G | p.Asn342Asp | rs267607482 | 8 | heterozygous |
| 33. | c.735+1G>T |  | rs397516698 | 2 | heterozygous |
| 34. | c.735+1del |  | n.a. | 2 | heterozygous |
| 35. | c.735+3A>G |  | rs267607483 | 2 | heterozygous |
| 36. | c.821T>G | p.Leu274Arg | rs267607494 | 4 | heterozygous |
| 37. | c.821T>C | p.Leu274Pro | rs267607494 | 6 | heterozygous |
| 38. | c.854C>T | p.Ala285Val | rs1368507241 | 1 | heterozygous |
| 39. | c.1009G>C | p.Ala337Pro | rs59962885 | 1 | heterozygous |
| 40. | c.1010C>T | p.Ala337Val | n.a. | 1 | heterozygous |
| 41. | c.1013T>G | p.Leu338Arg | rs57496341 | 1 | heterozygous |
| 42. | c.1034T>C | p.Leu345Pro | rs57639980 | 1 | heterozygous |
| 43. | c.1043A>C | p.Gln348Pro | rs1411703397 | 1 | heterozygous |
| 44. | c.1049G>C | p.Arg350Pro | rs57965306 | 7 | heterozygous |
| 45. | c.1064G>C | p.Arg355Pro | rs61368398 | 3 | heterozygous |
| 46. | c.1064G>A | p.Arg355Gln | rs61368398 | 1 | heterozygous |
| 47. | c.1076_1084del | p.Glu359_Ser361del | rs58409037 | 1 | heterozygous |
| 48. | c.1099A>T | p.Ile367Phe | rs62636494 | 10 | heterozygous |
| 49. | c.1109T>C | p.Leu370Pro | rs59308628 | 3 | heterozygous |
| 50. | c.1130T>C | p.Leu377Pro | rs1432061016 | 1 | heterozygous |
| 51. | c.1154T>C | p.Leu385Pro | rs57955682 | 1 | heterozygous |
| 52. | c.1166A>C | p.Gln389Pro | rs121913004 | 1 | heterozygous |
| 53. | c.1175T>C | p.Leu392Pro | rs62636493 | 1 | heterozygous |
| 54. | c.1203G>C | p.Glu401Asp | rs2125168897 | 21 | heterozygous |
| 55. | c.1205T>C | p.Ile402Thr | rs1553603571 | 2 | heterozygous |
| 56. | c.1216C>T | p.Arg406Trp | rs121913003 | 10 | heterozygous |
| 57. | c.1217G>A | p.Arg406Gln | rs1057520275 | 1 | heterozygous |
| 58. | c.1228G>A | p.Glu410Lys | rs2125168940 | 3 | heterozygous |
| 59.^**^ | c.1285C>T | p.Arg429* | rs150974575 | 1 | compound heterozygous |
| 60.^**^ | c.1288+1G>A |  | rs112224037 | 1 | compound heterozygous |
| 61.^**^ | c.1289-2A>G |  | rs398122940 | 1 | homozygous |
| 62.^**^ | c.1289-741G>A |  | rs985650778 | 2 | compound heterozygous |
| 63. | c.1237G>A | p.Glu413Lys | rs61726467 | 3 | heterozygous |
| 64. | c.1237_1238delinsCG | p.Glu413Arg | n.a. | 2 | heterozygous |
| 65. | c.1243_1244delCGinsGA | p.Arg415Glu | n.a. | 3 | heterozygous |
| 66. | c.1255C>T | p.Pro419Ser | rs62635763 | 7 | heterozygous |
| 67. | c.1325C>T | p.Thr442Ile | rs121913005 | 3 | heterozygous |
| 68. | c.1333A>G | p.Thr445Ala | rs267607498 | 1 | heterozygous |
| 69. | c.1358C>T | p.Thr453Ile | rs267607488 | 1 | heterozygous |
| 70. | c.1360C>T | p.Arg454Trp | rs267607490 | 10 | heterozygous |
| 71.^**^ | c.1366G>T | p.Gly456Trp | n.a. | 1 | homozygous |
| 72.^**^ | c.1366G>A | p.Gly456Arg | rs397516690 | 1 | homozygous |
| 73. | c.1370A>T | p.Glu457Val | rs267607496 | 5 | heterozygous |
| 74. | c.1379G>T | p.Ser460Ile | rs267607491 | 1 | heterozygous |

^*^All variants were re-adjudicated based on the NM_001927.4 transcript.

^**^ These variants were considered P/LP in the compound heterozygous or homozygous state, as reported. Their role in disease in heterozygous state is uncertain.

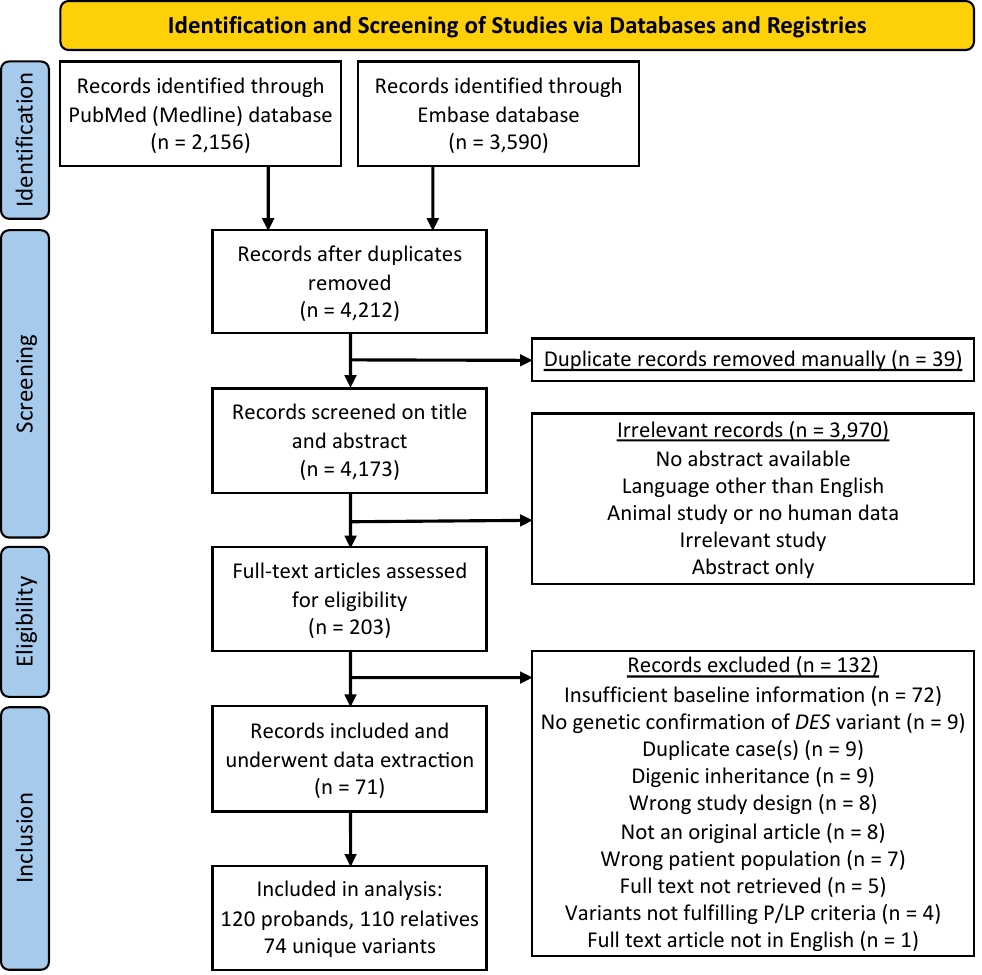

### Figure S1. Preferred Reporting Items for Systematic Reviews and Meta-Analyses (PRISMA) flow diagram.

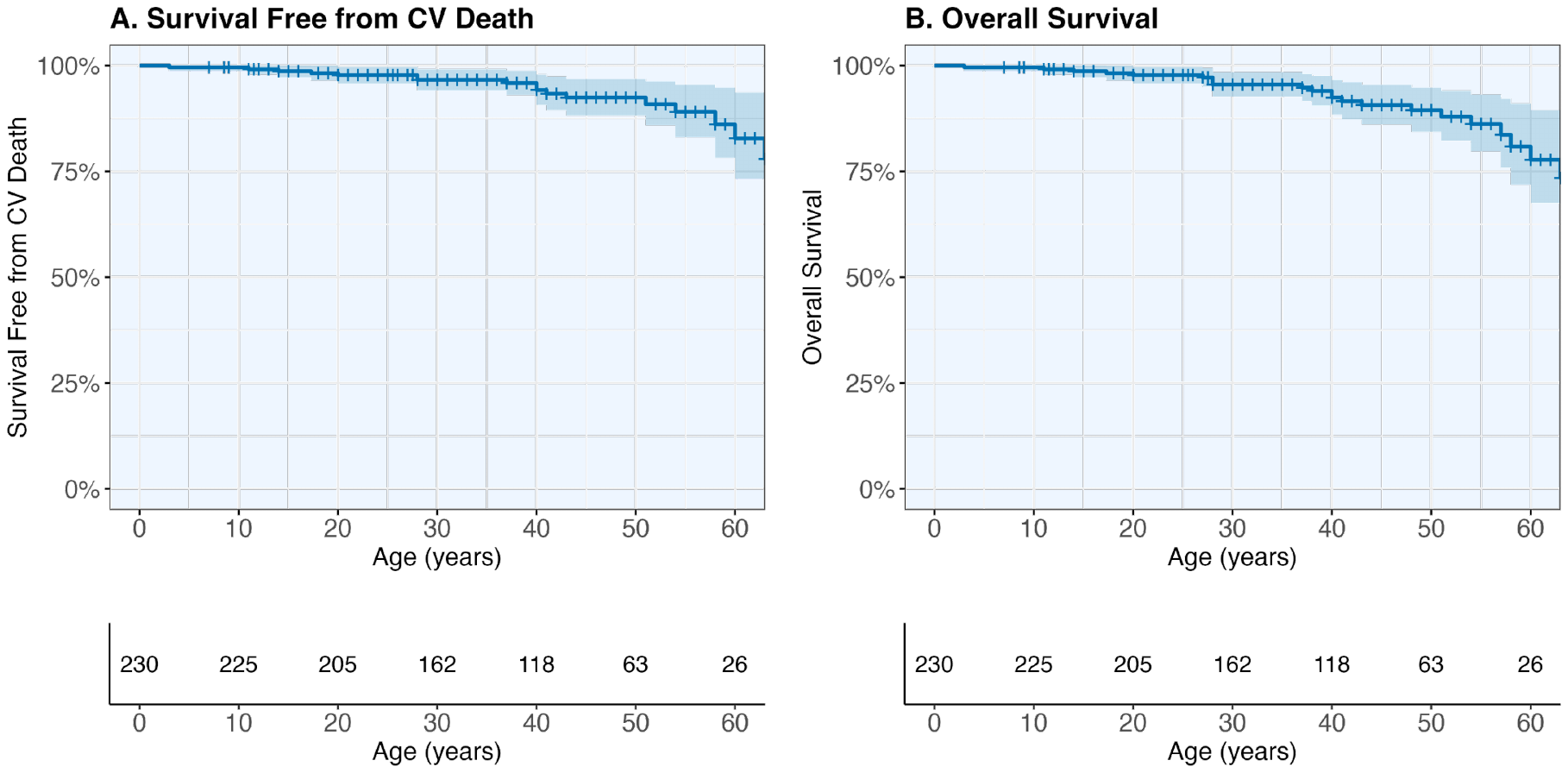

### Figure S2. Survival free from cardiovascular death and overall survival.

Survival free from cardiovascular (CV) death (**A**) and overall survival (**B**) are presented with 95% CIs (shaded area) in the overall population.
